# Verification of two Alternative Do-it-yourself Equipment Respirators Seal as COVID-19 Protection (VADERS-CoV): a quality assessment pilot study

**DOI:** 10.1101/2020.05.23.20111054

**Authors:** Marco Pettinger, Mona Momeni, Clemence Michaud, Michel Van Dyck, David Kahn, Guillaume Lemaire

## Abstract

**Background:** During the ongoing COVID-19 pandemic, healthcare workers are facing shortage in personal protective equipment, especially adequate respirators. Alternative do-it-yourself respirators (ADR) emerge, without any proof of protection.

**Objective:** Verify seal potential of two ADR compared to a common FFP2 respirator.

**Design:** Quality assessment pilot study.

**Setting:** Tertiary Care Hospital.

**Participants:** Ten anaesthesiology residents.

**Interventions:** Participants performed quantitative face-fit tests (QNFT) with three respirators to evaluate seal. A common FFP2 respirator was used as baseline (control group). ADR tested in this study are an Anaesthesia Face Mask (AFM) and a full-face Modified Snorkelling Mask (MSM) with a 3D-printed connector, both in conjunction with a breathing system filter.

**Main outcome measures:** Non-inferior seal performance of ADR over FFP2, assessed by calculated QNFT based on measured individual fit factors, as defined by the Occupational Safety and Health Administration.

**Results:** For each respirator a total of 90 individual fit factor measurements were taken. Within the control group, seal failed in 37 (41%) measurements but only in 10 (11%) within the AFM group and in 6 (7%) within the MSM group (*P* < 0.001 respectively). However, when calculating the final, mean QNFT results, no statistically significant difference was found between respirators. Successful QNFT were determined for 5 out of 10 participants in the control group, for 8 in the AFM group (*P* = 0.25) and for 7 in the MSM group (*P* = 0.69).

**Conclusion:** Both ADR do have the potential to provide non inferior seal compared to a common FFP2 respirator. While AFM respirators are easily assembled, snorkelling masks must undergo significant but feasible modifications. Our results suggest that those ADR masks might be further investigated as they seem to be viable alternatives for situations when certified respirators are not available.

**Trial registration:** Clinicaltrials.gov identifier: NCT04375774

## Introduction

To prevent transmission of the novel SARS-CoV-2 virus among exposed health care workers (HCW), the WHO recommends the use of tight-fitting facepiece respirators such as the FFP2 or N95.^1^ However, global personal protective equipment (PPE) stockpiles are shrinking rapidly because of the elevated demand during the COVID-19 pandemic and since the capacity to expand PPE production is limited while supply chains are disrupted and counterfeit deliveries discovered, severe local shortages of trustworthy respirators are a reality.^2-4^ Possible deconfinement measures are likely to increase demand even further as indications to wear respirators become broader (i.e. semi-urgent surgery for affected COVID-19 patients).

We hypothesized that this shortage of respirators may be counteracted by an Alternative Do-it-yourself Respirator (ADR) composed of widely available components in most hospitals. We therefore investigated whether a breathing system filter (BSF) plugged into an Anaesthesia Face Mask (AFM) held in place with a hook ring strapped to a silicone head harness could have the potential to act as a respirator providing EN-149 and EN-14683 protection levels.^5,6^ All proposed components are certified medical devices that are assembled without any modifications since they are compatible with one another. The unknown variables of this setup are filtration performance and overall airtightness. A high performance hydrophobic-coated BSF was chosen to avoid any leak from filter penetration. The main unknown variable of this AFM is airtightness in form of total inward leakage.

During the preparations of the study, reports emerged of HCW using full-face snorkelling masks as respirators.^7^ We had the opportunity to acquire a set of snorkelling masks and decided to include them in our study as they seem to be used as PPE without any proof of protection. Before using these snorkelling masks, significant modifications must be undertaken, depending on the model used, in order to seal the mask and fit a BSF. For this Modified Snorkelling Mask (MSM), the unknown variable is also total inward leakage which consists of possible leaks within the mask itself, between the non-standard connections and between the mask and face of the wearer.

The aim of this study was to verify seal performance of the AFM and the MSM with a validated evaluation method, being the quantitative fit test. We hypothesized that the two ADR have the potential to successfully pass individual and overall quantitative face-fit tests (QNFT) with non-inferior results compared to a common FFP2 tight-fitting face piece respirator. There are up to date no reported proofs of protection with these two ADR.

## Methods

Ethical approval for this study (Ethical Committee No. 2020/15AVR/226) was provided by the Ethical Committee of the Cliniques Universitaires Saint-Luc, Brussels, Belgium (Chairperson Pr. JM Maloteaux) on April 15 2020. The study was undertaken at the Anaesthesiology Department of the Cliniques Universitaires Saint-Luc, Brussels, Belgium from April 16 to 20 2020.

After giving informed consent, 10 healthy, volunteer anaesthesiology residents were included. Participants confirmed not to have smoked in the hour before testing and males were clean shaven within 12 hours. Participants performed QNFT with the three different respirators as defined by the Occupational Safety and Health Administration (OSHA).^8^ This test measures the concentration of particles inside and outside the respirator mask while participants perform predefined exercises. Any leak will be revealed by an increased concentration of particles inside the respirator mask. The ratio between these two concentrations is called fit factor.

Kim *et al*.^9^ questioned the reliability of the standardized exercises in reproducing head and body movements in a real-life healthcare worker (HCW) setting. Two additional exercises were therefore added to the conventional seven, to integrate routine HCW tasks which consisted of a video assisted intubation of a manikin with an Airtraq® (Prodol Meditec S.A, Vizcaya, Spain) and an ultrasound guided peripheral catheter placement on an i.v. phantom. The full test sequence of nine exercises for each respirator was composed as followed: normal breathing, deep breathing, head turning side to side, head moving up and down, talking, bending over, normal breathing, intubation, catheter placement.

During the test sequence of each respirator, nine individual fit factors were measured corresponding to the nine exercises, expressed as iFF. The harmonic mean of those nine iFF was calculated to determine the global fit factor (gFF) for the given participant and respirator, defining whether the QNFT was passed or not. The OSHA considers a QNFT for tight-fitting half facepieces like FFP2 respirators as passed when the gFF is equal to or greater than 100, even if some iFF lay below this threshold.^8^ The formula used to calculate the global fit factor is:

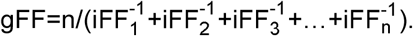

The device used in this study is the OSHA accepted PortaCount® Pro+ model 8038 (TSI® Inc, Shoreview, US) which has an integrated N95-Companion™, in conjunction with the model 8026 Particle Generator to guarantee a stable and sufficient ambient particle concentration. The principle of the PortaCount is based on a condensation particle counter. The concentration range of the PortaCount is from 0.01 to 500 000 particles/cm^3^ with a particle size range from 0.02 μm to greater than 1 μm.^10^ A fit factor of 200 is the highest number that can be displayed when using the N95-Companion. In practice we did not use the N95-Companion in order to assess plain raw data for every respirator model since the FFP2 respirator chosen had a high enough filtering performance to give reliable results without the N95-Companion.

QNFT were executed with the following respirators:

Control FFP2 respirator: a unisize duckbill FFP2 respirator in form of a model PFR P2, FILTERING HALF MASK FFP2 (O&M Halyard UK Ltd, Manchester, UK).

AFM respirator: size 4 or 5 autoclavable ClearFlex economy silicone anaesthetic mask with its hook ring and head harness (Intersurgical Ltd, Berkshire, UK) used in conjunction with BSF in form of a Gibeck® Iso-Gard® HEPA Light hydrophobic bacterial/viral filter (Teleflex Inc, Wayne, USA).

MSM respirator: size SM or ML Subea Easybreath© 500 full-face snorkelling mask (Decathlon SA, Villeneuve d’Ascq, France) used in conjunction with our own designed 3D-printed connector clipped onto the mask instead of the tuba piece to plug in the BSF. The chin valve has been sealed with tesa® extra Power Universal duct-tape (Tesa SE, Norderstedt, Germany). This water purge could act as an expiration valve to facilitate breathing and reduce fogging but the potential risk of it failing led us to the conclusion that it is safer to seal it. Since it has not been thoroughly tested for this use beforehand, like the ones found in some FFP2 models, its potential failure and staying in an “always open” position could lead to a major leak hazard.

**Figure 1.**
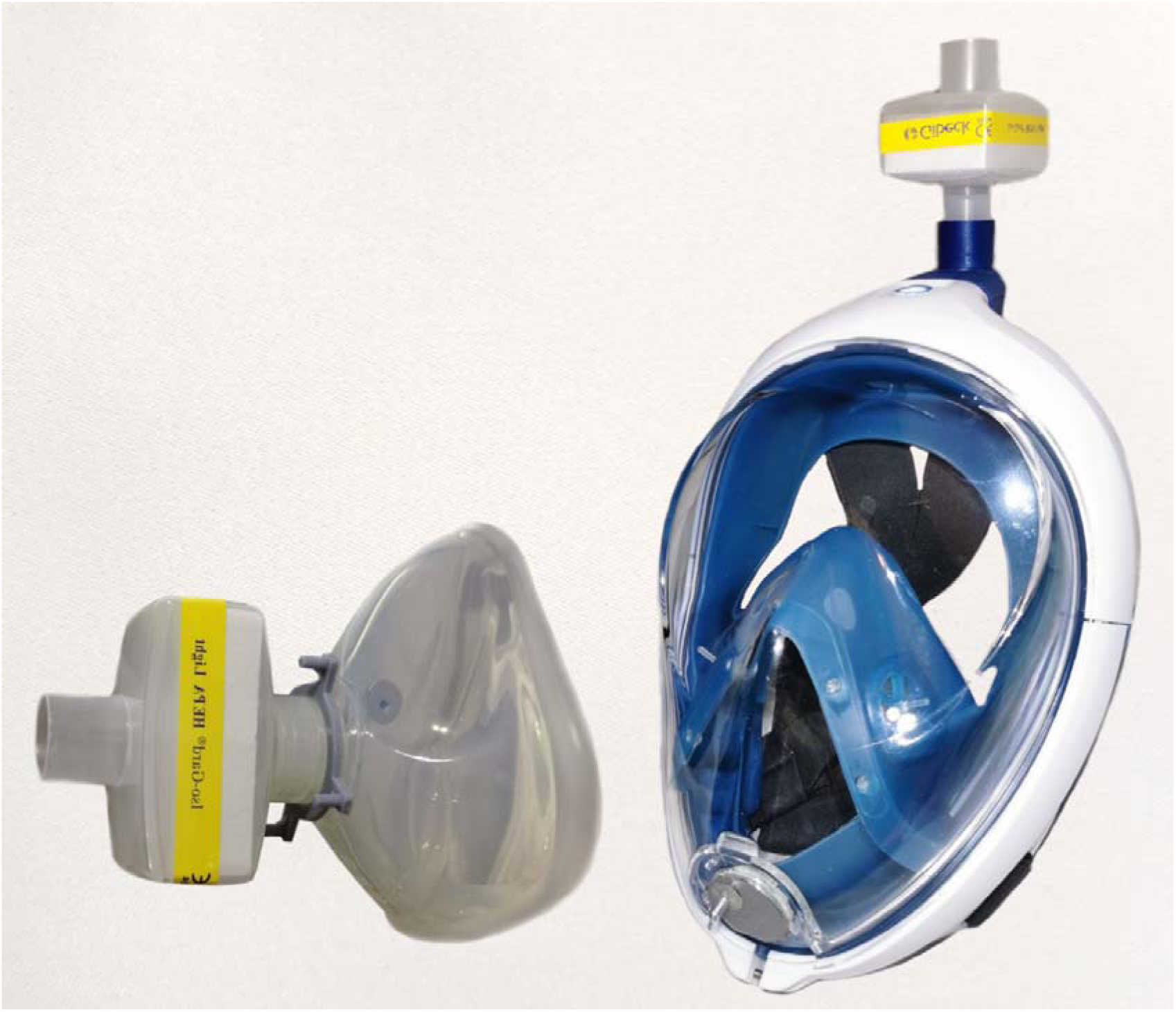
ADR as used for testing. Anaesthesia Face Mask on the left and Modified Snorkelling Mask on the right, both with filter and probe for the PortaCount in place.

The PortaCount probes for the FFP2 were installed following the instructions in the manual. A tubing support neck strap was used to prevent the weight of the probing tube affecting seal performance. Probes have been installed laterally in the AFM and through the sealed chin valve in the MSM to assure proximity to airflow and not risk being blocked by skin.

Subsequently, participants subjective feelings were surveyed for comfort, ease of breathing, field of view, ease of equipping and general appreciation for all tested respirators on a Likert scale (choices: bad, insufficient, acceptable, good). Free commentary was possible but optional.

FitPro+™ Fit Test Software (TSI® Inc, Shoreview, USA) was used to transfer data from the PortaCount device. Data was entered into Microsoft® Excel® for Office 365 MSO (Microsoft® Corp, Redmond, USA) for statistical evaluation. IBM SPSS® Statistics V.23 (IBM Corp, Armonk, US) was used for statistical analysis.

A sample size of 10 participants was chosen as we followed the European standard EN-149:2009-08 stating in section ‘7.9.1 Total inward leakage’ that it requires only 10 respirators to be leak tested for certification.^6^ Since the respirators must be altered irreversibly for fit testing and cannot be reused, we thought it to be unsuited to increase sample size beyond this number because our study was performed during the state of emergency and during the global scarcity of PPE. Further, only a sample size of nine should be enough to verify the supposed sufficient seal in FFP2 described by Ciotti *et al*.^11^ (18.3%) and a presumed success rate of 80% with ADR considering a power = 0.8 with a significance level of 0.05.

The exact McNemar’s test was used to compare successful fit factor results between groups. The statistical evaluation of the survey was performed with an exact sign test, considering the answers to the survey as ordered categorical data. A value of *P* < 0.05 was considered as statistically significant.

## Results

Ten participants (five females and males), were included in the study and all performed three QNFT sequences. A total of 90 iFF were measured resulting in 10 gFF for each respirator.

Measured iFF for each respirator are represented in Fig. 2. There was a significant difference in terms of seal potential between ADR groups and the control group when comparing their iFF below the threshold of 100. Seal based on iFF measurements failed in 37 (41%) out of 90 exercises within the control group, in 10 (11%) within the AFM group and in 6 (7%) within the MSM group. The difference in the proportion of iFF below 100 between the FFP2 and AFM on one hand and between the FFP2 and MSM on the other hand, was statistically significant (*P* < 0.001, respectively). There was no significant difference between the AFM and the MSM group (*P* = 0.34).

**Figure 2.**
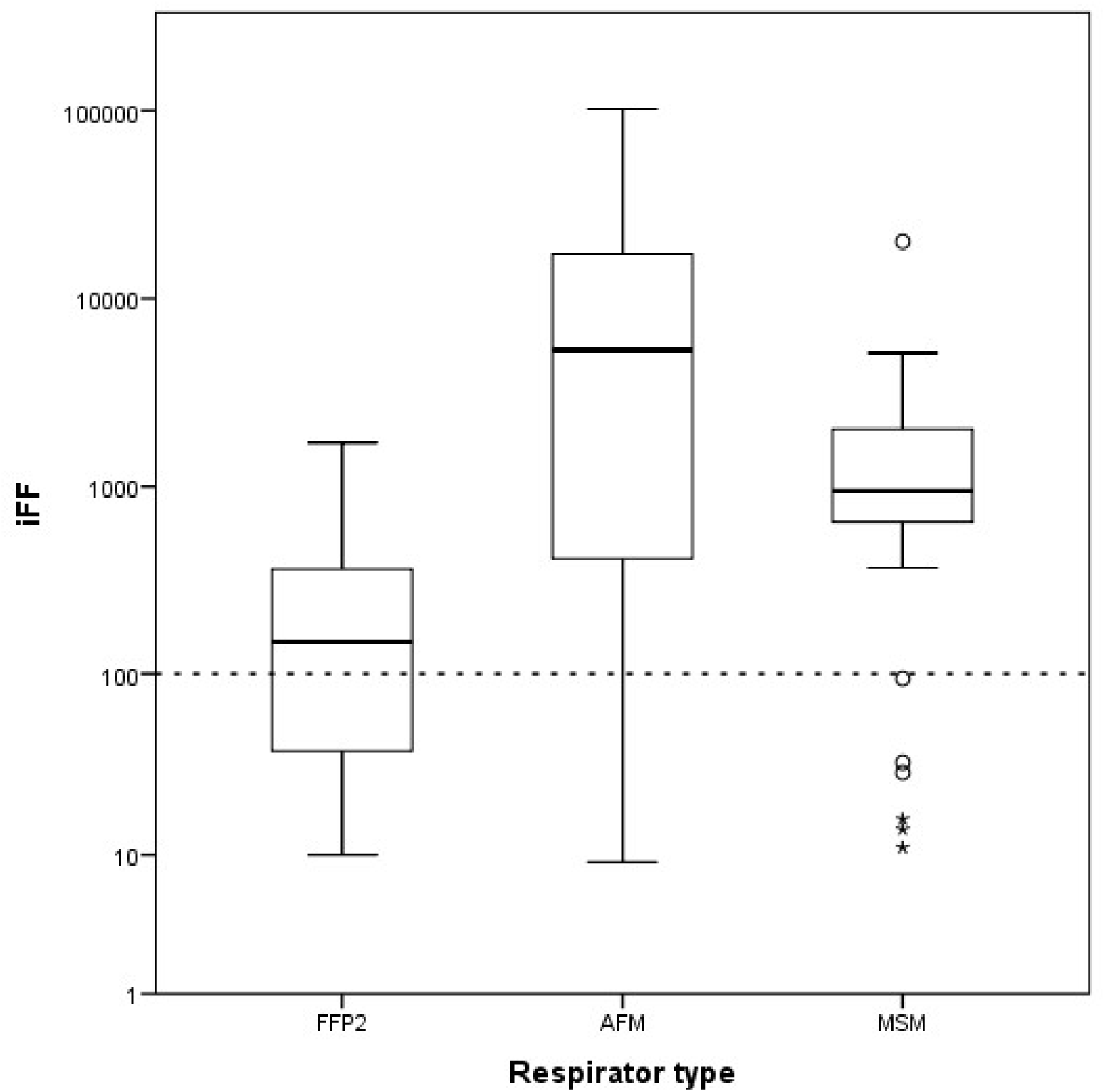
Box and whisker plot representation of raw iFF measurements for each respirator, logarithmic scale. The dotted line corresponds to a fit factor of 100. The solid line represents the median, the box represents interquartile range and whiskers represent maximum and minimum values without outliners, hollow circles represent outliners and asterisks represent extremes.

To ease comparison between the different respirators, we capped the maximum iFF values at 200 for all respirators before calculating gFF, since this is the conventional limit from the PortaCount N95-Companion for tight-fitting half facepieces. This cap also offers a more conservative evaluation as a too low iFF in one exercise cannot be compensated by several high iFF when calculating the gFF, even though the harmonic mean already strongly tends towards the lowest measurement.^12^ As stated before, a QNFT is passed if the gFF is greater than or equal 100. There was no statistically significant difference between each ADR and the control group when comparing QNFT results. Successful QNFT were determined in 5 out of 10 within the control group compared to 8 in the AFM group (*P* = 0.25) and compared to 7 in the MSM group (*P* = 0.69). There was no difference between both ADR in passing QNFT (*P* = 1.00).

All these evaluations have been reanalysed without the two specific HCW tasks, considering only the first seven conventional exercises as defined by the OSHA. In this setup, 26 out of 70 (37%) exercises measured iFF below 100 in the FFP2 control group. This was the case in 9 out of 70 (13%) exercises within the AFM group and in 5 out of 70 (7%) exercises within the MSM group. Successful QNFT results following gFF calculation increased from 5 out of 10 in the 9-exercises FFP2 group to 7 out of 10 in the 7-exercises group (P = 0.69). No changes between each of the ADR groups were observed.

The results of the survey are represented in Fig. 3. Vision was evaluated significantly impaired with the MSM respirator compared to the FFP2 (*P* = 0.031). No significant difference was found in the survey between the FFP2 and AFM respirator. In the free commentary, the most cited problems were communication difficulties for both ADR, the difficulty to combine with glasses regarding the AFM and the impossibility to combine with glasses while using the MSM.

**Figure 3.**
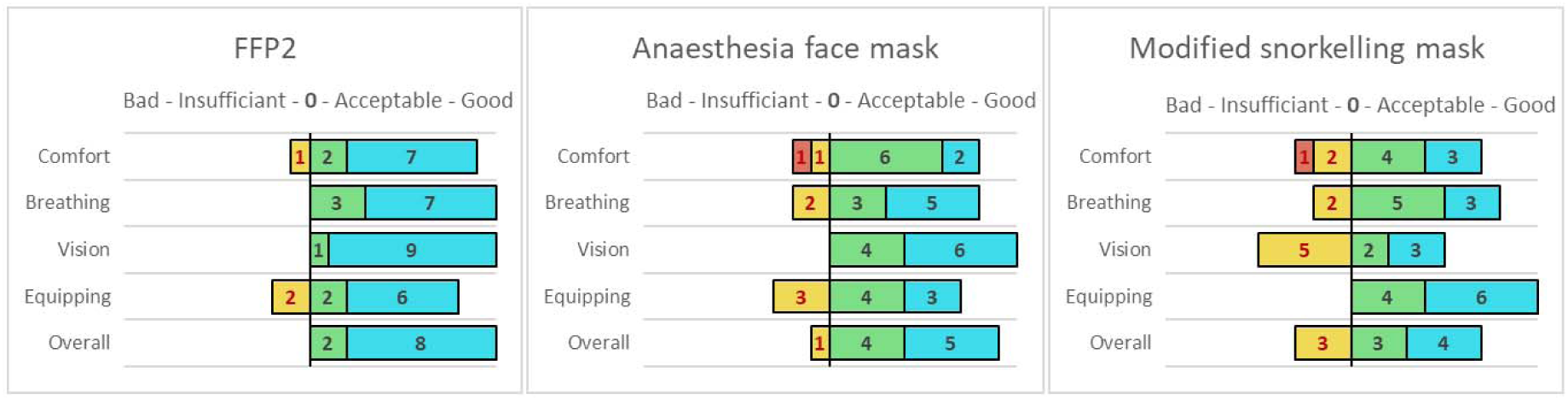
Diverging stacked bar chart representation of survey results for each respirator type.

## Discussion

The current study confirms that the tested ADR have the potential to pass fit tests in the first place. The results of the ADR performances are intriguing as they have the potential to achieve confirmed fit factor values of up to several thousand. While providing high iFF is interesting, remaining above the threshold of 100 is truly decisive and we observed several measurements below this value. We explain their momentary drops below 100 due to their rigid structure, limiting their capacity to adapt to facial movements. This happened in 10 out of 90 cases within the AFM group and in 7 out of 90 cases within the MSM group. However, it was significantly less frequent than in the FFP2 control group where this was the case in 37 out of 90 cases. When transforming iFF into overall gFF, to determine whether QNFT was passed, the difference between ADR and FFP2 was no longer statistically significant. We believe this is due to the very conservative calculations concerning gFF, as explained earlier, but also because our pilot study’s sample size was too small, since our FFP2 performed better than expected.^11^ There is no significant difference between the AFM and MSM respirators since both perform equally in terms of iFF measurements and QNFT results.

We observed an increase in successful QNFT with the FFP2 respirator from 5 to 7 out of 10 when removing the additional two HCW tasks from the test sequence. The added tasks during testing did not significantly affect outcomes but the rise in successful QNFT results tend to confirm Kim *et al*.^9^ findings that the proposed standard sequence seems to be insufficient to assess fit during a variety of real-life head movements.

For one participant, none of the tested respirators had a successful QNFT, underlining the importance of preliminary fit testing in order to find an adapted model. As a matter of fact, yearly fit testing is a basic security routine in the USA and enforced by law since 1976.^13^ To this date, this is not the case in Europe as the non-binding 89/656/EEC European directive on the minimum health and safety requirements barely mentions face fit testing. Some European countries have drafted their own laws, like the UK where fit testing is mandatory when respirators are required.^14^ Noti *et al*.^15^ showed however that a non-fitted respirator does not provide better protection than a loose surgical mask.

The existence of this security gap is supported by Ciotti *et al*.,^11^ who found that FFP2 duckbill models were adapted to the facial morphology of HCW in only 18.3% of the cases and in 57.5% with cardboard models. Even though the QNFT pass rate in our FFP2 duckbill control group was higher than anticipated, unfitted respirators do not provide the expected seal levels.

Concerning the survey, both ADR were less popular among participants than the common FFP2 and vision was evaluated as significantly impaired compared to the FFP2. First, it is not possible to wear glasses in combination with this mask. Second, there is a rapid formation of fog from condensation even though we did not alter the designers fog reducing “one way” air flow principle, since we kept the internal valves. This might further increase with wearing time beyond the duration of our test. Third, the design of the viewing glass itself reduces the peripheral field of view. Participants reported difficult breathing with the MSM, and some found it oppressing to wear a full-face mask.

Conveniently combining the AFM with glasses was sometimes challenging depending on the model, especially with the size 5, as it covers the nose completely. The head harness’s straps might be used to help them stay in place but cannot replace a secure positioning on the nose. Overall, the quite voluminous BSF did not majorly impair vision.

Communication was impacted with both ADR as comprehension of the wearer was impaired. This should not be underestimated as it might alter workflow and even patient security when confronted to a critical situation in the operating theatre for example.

The adequate BSF must be chosen carefully to fulfil the required norms as their performances vary.^16^ Wilkes tested a total of 33 different BSF for their filtration performance and found important differences between manufacturers and models: 14 of 24 electrostatic filters did not comply with N95 norms but all of the 9 hydrophobic filters complied with at least N99 norms.^17^

The AFM respirator assembly is quite straight forward, takes seconds to be completed and only requires generally available components in hospitals. In practice, the head harness could be replaced by anything providing sufficient hold, from a rubber tourniquet to a gauze dressing, and a missing hook-ring could easily be 3D printed.

The fabrication of the MSM respirator is however more complicated since substantial but feasible modifications must be undertaken before it can possibly be used as a respirator. None of those fulfil any security norms, nor has any certification organism approved them. If a water purge is present, its valve should be removed and sealed shut. The tuba piece should be removed to facilitate designing a tight-fitting 3D-printed connector for the BSF.

Cost is a minor factor in these setups, as the major parts are reusable after disinfection. Running costs concern the BSF and the 3D-printed connector. BSF with the highest filtering classes have similar prices as FFP2 respirators in our institution, but prices might differ in other regions.

The study has limitations that need to be acknowledged. The study methodology was designed to approximate EN-149 European standardization protocols that certify FFP2 respirators, which only need 10 respirators to be tested.^5^ However, the sample size of 10 subjects is small for any statistical interpretation and also represents only a limited variety of facial morphologies. We did not take face measures but included participants of both sexes equally to enforce a certain variety in facial morphologies. As stated in the PortaCount manual, it should be clarified that the measurement provided by the PortaCount is an assessment of respirator fit during the fit test only. Respirator fit at other times will vary. The fit factor value is not intended for use in calculating an individual’s actual exposure to hazardous substances.^10^ This study does not fulfil any certification norms and should not be considered as an approval to use noncertified PPE. Extrapolating the results presented in this study to other individuals or materials than those tested is up to the reader’s discretion. Certified and ideally preliminary face fitted respiratory PPE should always be used when available. Proof of airtightness alone is no proof of real-life protection from hazardous aerosols. We surveyed for several subjective comfort evaluations, but participants wore the different respirators only for several minutes which does not reflect a realistic wearing duration. However, the current state of emergency and potential lack of certified PPE inspired us to realize this study, to give HCW some clarity on the matter in order to aid them in their own risk assessment, when being forced to look for alternatives.

In conclusion, the tested ADR do have the potential to provide non inferior seal performance than a common FFP2 respirator as verified by quantitative fit testing. While anaesthesia face masks are easily assembled with available components in hospitals, snorkelling masks have to be acquired additionally and must undergo carefully executed, significant modifications. Special attention needs to be paid when choosing an adequate filter. Our results suggest that those alternative respirators should be further investigated, as they seem to be viable alternatives for situations when certified PPE is not available. Individual face fit testing should be used to guide HCW in choosing the respirator best adapted to their facial morphology.

## Data Availability

Not yet published.

## Acknowledgements relating to this article

Assistance with the study: We thank Vandeputte Safety Experts Belgium for their helpful technical insight and for providing the PortaCount device free of charge and Decathlon Belgium SA for providing the two snorkelling masks free of charge. We thank all the participants and the Department of Anaesthesia, Cliniques Universitaires Saint-Luc, Brussels for their sincere support and insight during this study. We specially thank Thomas Schubert M.D. Ph.D., Florent Dumonceaux M.D. and Arthur Bun M.D. for their practical help during this study.

## Financial support and sponsorship

All remaining costs were covered by the Department of Anaesthesia, Cliniques Universitaires Saint-Luc, Brussels. This research did not receive any specific grant from funding agencies in the public, commercial, or not-for-profit sectors.

## Conflicts of interest

none

## Presentation

none

## Notes

### Competing Interest Statement

The authors have declared no competing interest.

### Clinical Trial

NCT04375774

